# Age-severity matched cytokine profiling reveals specific signatures in Covid-19 patients

**DOI:** 10.1101/2020.07.28.20162735

**Authors:** Roberta Angioni, Ricardo Sanchez-Rodriguez, Fabio Munari, Nicole Bertoldi, Diletta Arcidiacono, Silvia Cavinato, Davide Marturano, Annamaria Cattelan, Antonella Viola, Barbara Molon

## Abstract

A global effort is currently undertaken to restrain the COVID-19 pandemic. Host immunity has come out as a determinant for COVID-19 clinical outcome, and several studies investigated the immune profiling of SARS-CoV-2 infected people to properly direct the clinical management of the disease. Thus, lymphopenia, T-cell exhaustion, and the increased levels of inflammatory mediators have been described in COVID-19 patients, in particular in severe cases^1^. Age represents a key factor in COVID-19 morbidity and mortality^2^. Understanding age-associated immune signatures of patients is therefore important to identify preventive and therapeutic strategies. In this study, we investigated the immune profile of COVID-19 hospitalized patients identifying a distinctive age-dependent immune signature associated with disease severity. Indeed, defined circulating factors - CXCL8, IL-10, IL-15, IL-27 and TNF-α - positively correlate with older age, longer hospitalization, and a more severe form of the disease and may thus represent the leading signature in critical COVID-19 patients.

---

We evaluated the hospitalization time (HT), immune and clinical features in a cohort of 44 *SARS*-*CoV*-*2*-positive symptomatic patients who have been admitted at the University Hospital of Padova from 9.04.2020 to 5.05.2020. The demographic and clinical data of patients are reported in Table 1. We selected three parameters that may be relevant to stratify COVID-19 patients, such as patients’ age, HT and disease severity (DS) as defined by the WHO guidelines^3^.

**Table 1.**
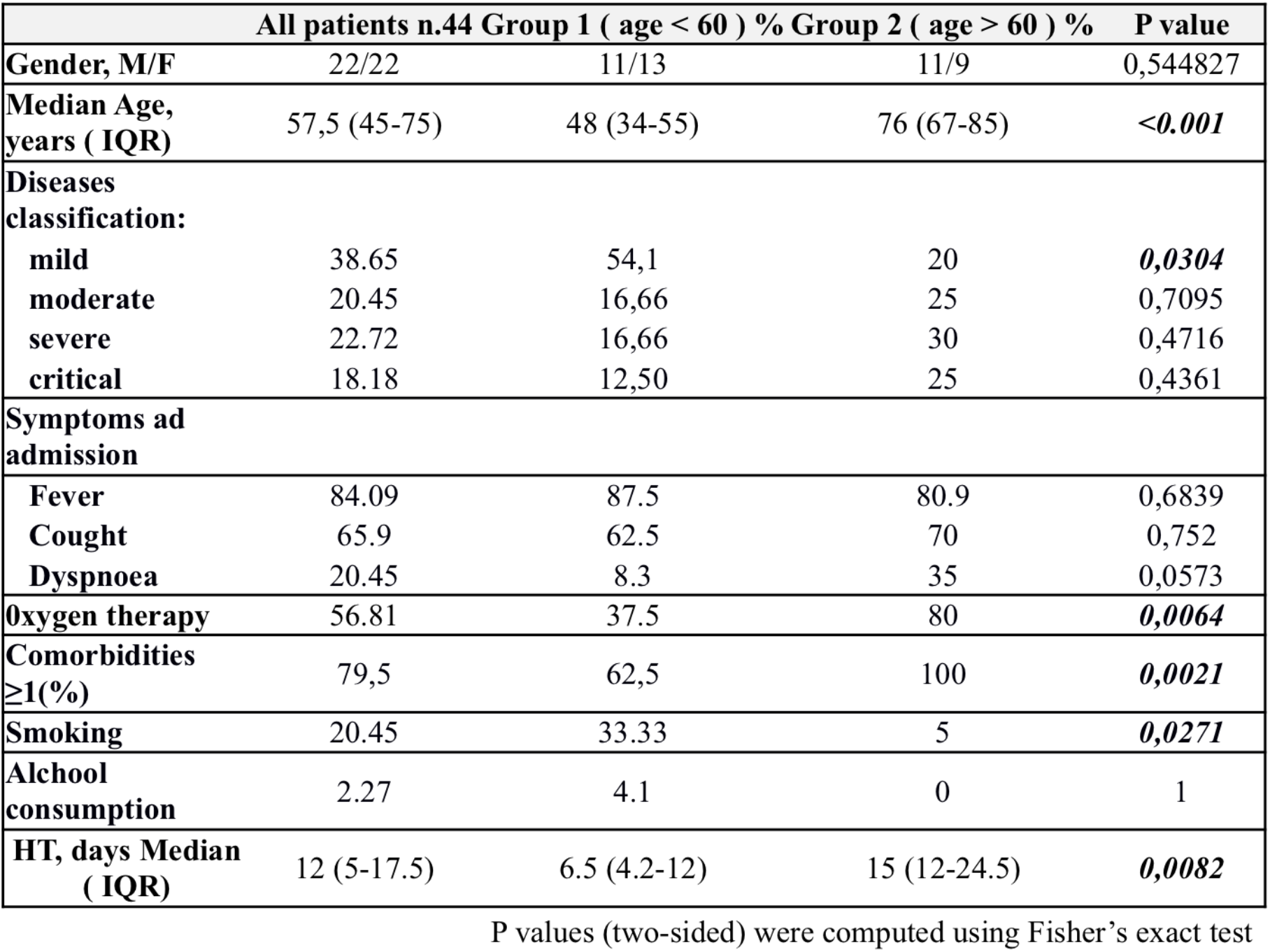
Demographic and Clinical data of COVID19 patients at the HA.

By performing correlation analysis in our clinical dataset, we obtained a significant positive correlation between age and HT (R Pearson 0.35350 Fig1A) and, in agreement with the current literature^4,5^, we confirmed a positive association between age and DS (R Pearson 0.4445, Fig1B), and HT versus DS (R Pearson 0.6568, Fig1C) in our cohort.

The generation of IgG antibodies against SARS-CoV-2 proteins might represent an applicable parameter for COVID-19 patient stratification. Nevertheless, the parallel between SARS-CoV2 seropositivity and the clinical outcome is still matter of investigation^6,7^. In our cohort, 34% of the patients showed positive IgG titer against the SARS-CoV-2 spike protein at the admission time (AT) (Fig1 SA). However, no correlation between IgG positivity and HT (Fig 1S B), age (Fig 1S C), or DS (Fig 1S D) was evident.

**Figure 1.**
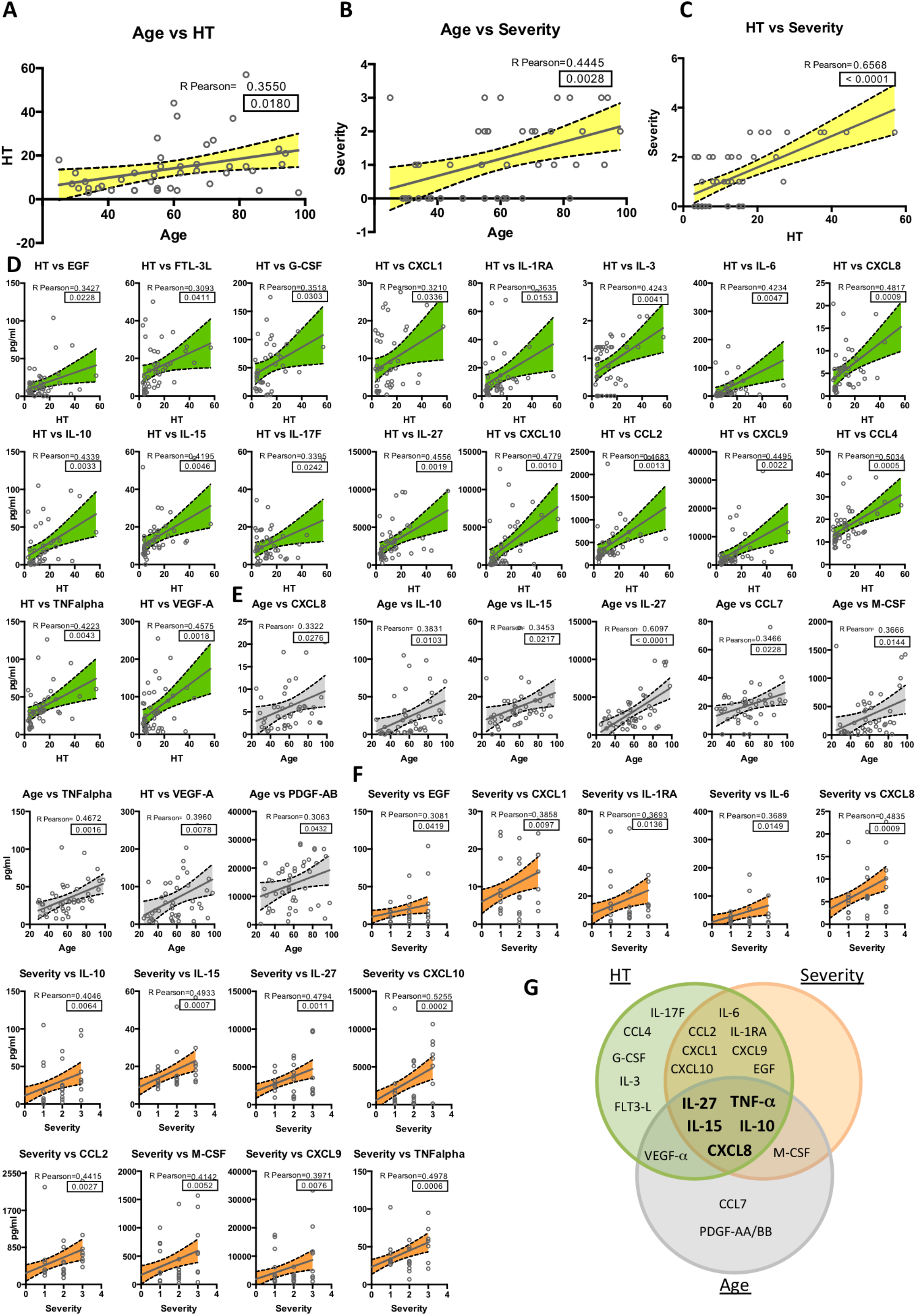
Correlation Analysis in COVID-19 patient cohort. Positive correlation between age and HT **(A)**, age and DS **(B)** HT and DS **(C)** was measured by Person coefficient r (95% confidence interval) and two-tailed p-value analysis (indicated inside the square). Positive correlation between HT **(D)**, age **(E)** and DS **(F)** and the indicated analytes was measured by Person coefficient r (95% confidence interval) and two-tailed p-value analysis (p-value is indicated in the square). **G)** Venn diagram showing cytokines, chemokines, and growth factors as related to Age (*Gray*), HT (*Green*) and DS (*Orange*).

Although it has been reported that COVID-19 mortality is higher in men than in women^8^, we did not observe major differences in the HT between females (50%) and males (50%) in our cohort study, with an HT mean of days 9,2 +/- 1,68 for men and days 7,7 +/- 1,4 for women (Fig 1S E) and not even a significant variation in the timing from symptom onset to hospital admission (Fig 1S F) and hospital discharge (Fig 1S G).

A properly-coordinated immune response represents a mandatory requirement for the clearance of SARS-CoV-2 infection^9^. Importantly, circulating factors play a crucial role in the immunopathology of SARS-CoV-2 infection and, in some cases, they might also tailor patient clinical path^10^. To outline the prevailing immune milieu in our cohort, we quantified cytokines and growth factors in patients’ plasma at admission time. To this aim, by multiplexed analysis, we concomitantly measured 48 circulating analytes and we performed correlation analysis between the plasma concentration of each analyte and HT, age, or DS. Interestingly, we identified a distinctive pattern of cytokines showing positive correlation with either HT (Fig 1D), age (Fig 1E), or DS (Fig. 1F). On the other side, an additional set of cytokines unveiled no association with HT (Fig 2S), age (Fig 3S) or DS (Fig 4S). The Venn diagram represents unique set of cytokines that were differentially expressed in the three groups (Fig 1G). As expected, the cytokine signatures associated with HT and DS were partially overlapping (12 out of 18 for HT, 12 out of 13 for DS). These shared cytokines include molecules that have been implicated in COVID-19 pathogenesis such as IL-1RA^11^, IL-6, CXCL10, CXCL8, IL-10, CCL2, CXCL9 and TNF-α^12,13^, as well as molecules that have not been associated to severity yet, such as IL-15, IL-27 and EGF. Interestingly, a specific subset of nine factors was selectively increased in older patients. Among these, we singled out a unique set of five cytokines - CXCL8, IL-10, IL-15, IL-27 and TNF-α - shared among the three variables. We also pointed out a defined cytokine trait (IL-6, CXCL9, IL- 1RA, CXCL1, CXCL10, CCL2, EGF) of more severe COVID-19 patients, which is independent from age.

**Figure 2.**
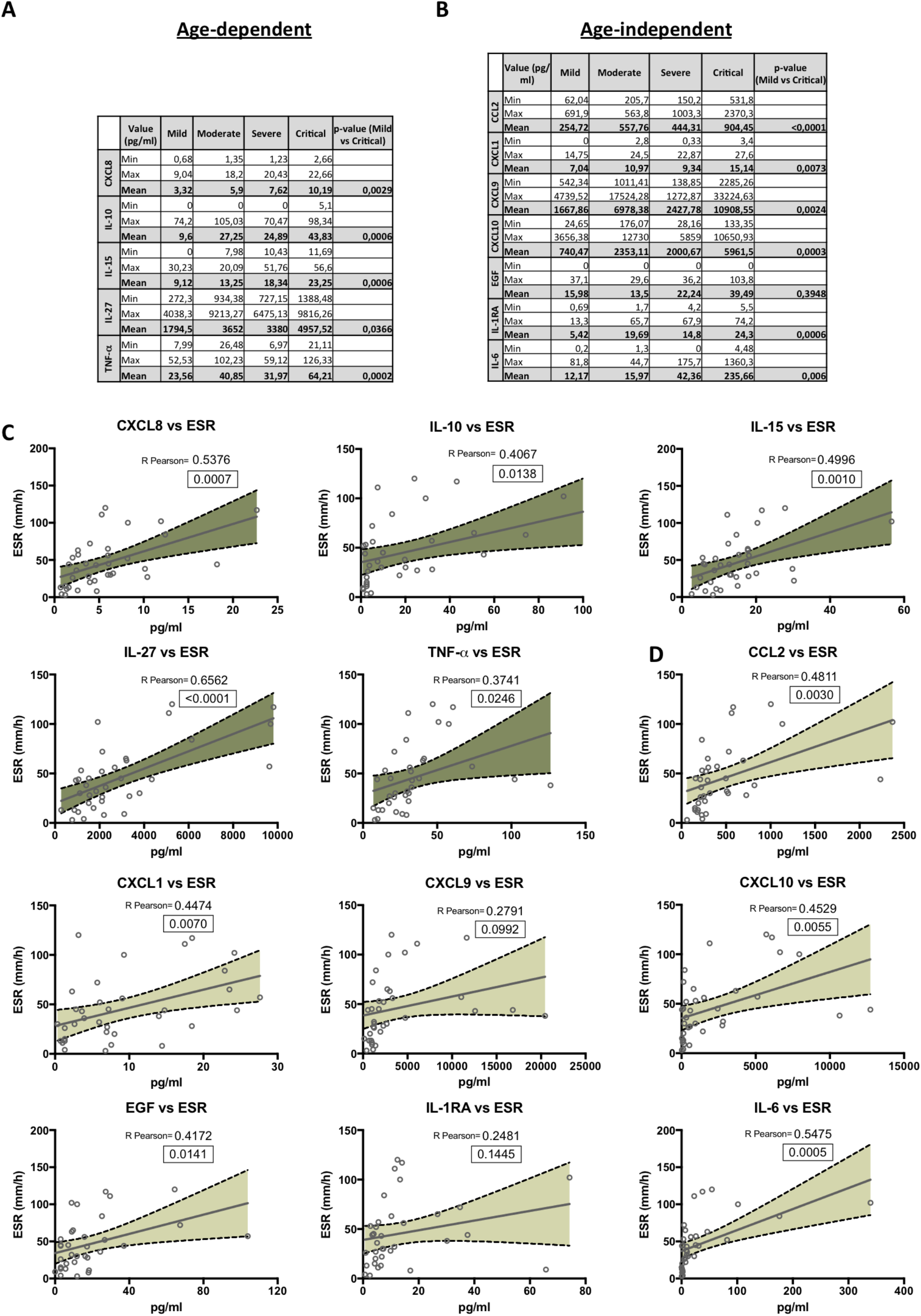
Clinical relevance of COVID-19-related cytokines. The minimum (Min), maximal (Max) and mean plasma concentration values (pg/ml) of age-dependent **(A)** or age-independent **(B)** cytokines that correlate with both HT and DS are indicated for patients with mild, moderate, severe or critical clinical score. The P value (mild versus critical) has been calculated using non-parametric Mann-Whitney test. Positive correlation between age-dependent (**C**) or age-independent (**D**) cytokines and ESR was measured by Person coefficient r (95% confidence interval) and two-tailed p-value analysis (indicated inside the square).

**Figure 3.**
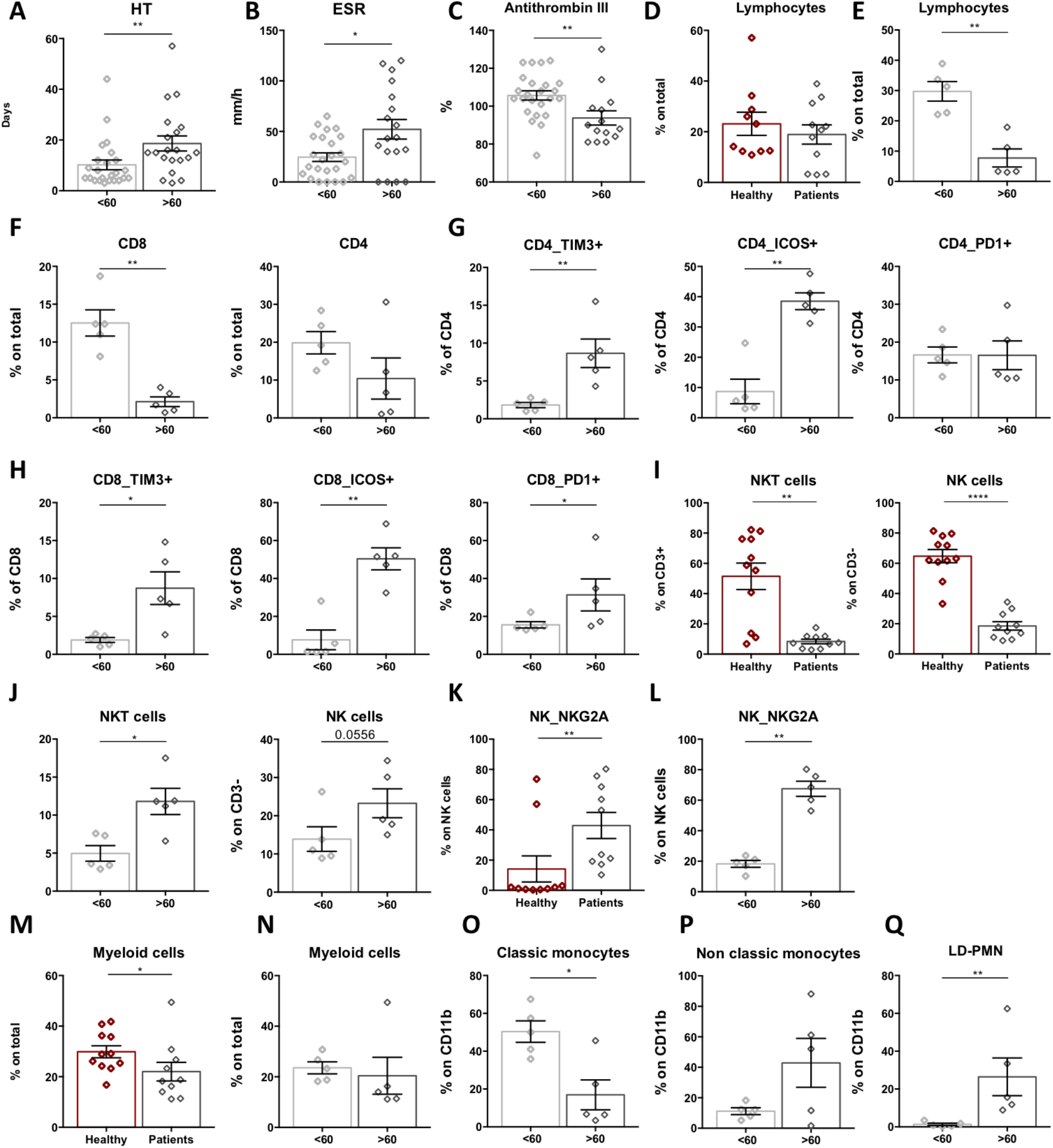
Age-dependent immune profiling of COVID-19 patients. Bar-charts showing HT, days **(A)**, ESR, mm/h (**B**) and antithrombin III, % (**C**) in younger (<60) or older (>60) patients. Bar-charts showing T lymphocyte percentage (CD3^+^ T cells) in healthy, age-matched controls and COVID-19 patients (**D**), or in COVID-19 patients grouped by age (**E**). FACS analysis of immune cell subsets in younger (<60) and older (>60) COVID-19 patients: percentage of CD8^+^ or CD4^+^T lymphocytes (**F**); expression of TIM3, ICOS and PD1 in CD4^+^ (**G**) or CD8^+^ T cells (**H**); percentage of NKT and NK cells in healthy age-matched controls and COVID-19 patients (**I**); percentage of NKT and NK cells in younger (<60) and older (>60) COVID-19 patients (**J**); NKG2A expression in NK cells in healthy age-matched controls or COVID-19 patients (**K)** and in younger (<60) or older (>60) COVID-19 patients (**L**); percentage of CD11b^+^ myeloid cells in healthy age-matched controls and COVID-19 patients (**M**) and in younger (<60) or older (>60) COVID19 patients (**N**); percentage of classical CD11b^+^CD14^high^CD16^-^ (**O**), and non-classical CD11b^+^CD14^low^CD16^+^ (**P**) monocytes, and LD-PMN CD11b^+^HLA-DR^lowneg^CD14^-^CD15^+^CD66b^+^(**Q**) in younger (<60) or older (>60) COVID-19 patients. Data are represented as mean of percentage of parent population ± SEM. Gating strategy in Fig S6. Mann-Whitney test; * p<0.05, ** p<0.01, ****p<0.0001.

To further associate the identified cytokine profiles to the clinical evaluation, the absolute plasma concentrations of the aforementioned age-dependent and age-independent cytokines were compared in patients with different disease severity, confirming that the cytokines were significantly up-regulated in critical cases, as compared to patients with mild disease (Fig 2A-B). In addition, the circulating levels of these cytokines positively correlated with diagnostic parameters such as ESR (Fig 2D-E), CPR and fibrinogen concentration (Fig 5S).

Patients’ age has been appointed as a crucial determinant for the response to SARS-CoV-2, being older people generally at higher risk of severe illness^14,15^. In addition, it has been described that 80% and 90% of deaths have occurred in patients aged >70 years and >60 years in Korea and Italy, respectively ^16^.In line with this evidence, our results identified a specific cytokine profile associated to COVID-19 in older patients. To further characterize patients’ immune responses in relation age, we performed a detailed characterization of circulating immune cells in a subset of patients at the admission time (Fig 3 and Fig 10S), by performing multiparametric FACS analysis of peripheral blood cells. We stratified our cohort study by fixing the age of 60 as the cut-off for patient grouping. Of note, these 2 groups clearly differed in terms of clinical parameters, with age >60 patients having longer hospital stay (18.65± 2.96 S.E.M) as compared to the younger (10.21 ± 1.92 S.E.M) (Fig3A), higher ESR (Fig.3B) and lower antithrombin III (Fig. 3C) at the admission time. Moreover, although we did not observe divergences in the total CD3^+^ lymphocyte counts in COVID-19 patients as compared to age-matched controls (fig 3D), a significant reduction in lymphocyte number appeared when our patients were stratified in >60< years old (Fig.3E).

The flow cytometry analysis of peripheral blood cells showed that the reduction in total CD3^+^ T cells observed in older patients was retained in the CD8^+^ T cell subsets, with a similar trend in the CD4^+^ compartment (Fig.3F). Additionally, in patients over 60 years, the level of expression of TIM3, ICOS and PD1 molecules dramatically increased on both CD4^+^ (Fig. 3G) and CD8^+^ T lymphocytes (Fig. 3H), suggesting that T cells are characterized by an inactive-exhausted phenotype in this group. Consistently with previous reports^9^, we confirmed a general decrease in the count of NK T and also NK cells in patients, when compared to healthy subjects (Fig. 3I). However, but very interestingly, we detected an increased number of NK T cells in the >60 patients compared to the <60 ones, with a similar trend in the NK compartment (Fig 3J.). As previously reported^17^, NK cells in COVID-19 patients presented an exhausted phenotype, as defined by the expression of the higher expression of the inhibitory receptor NK group 2 member A (NKG2A), compared to healthy donors (Fig. 3K); but, remarkably, this higher NKG2A expression was specific of the over 60 group (Fig.3L). Finally, we detected a slight but significant reduction of the total CD11b^+^ events in patients compared to controls, but no age-related differences were observed (Fig. 3M). Age-dependent differences were observed in terms of monocyte phenotype, being the classical monocytes (CD11b^+^/CD14^high^/CD16^-^) less represented in >60 patients as compared to the non-classical monocyte (CD11b^+^/CD14^low^/CD16^+^ subset) (Fig.3M). Moreover, we observed an expansion of a subset of granulocytic cells (CD11b^+^/HLA-DR^lowneg^/CD14^-^/CD15^+^/CD66b^+^) in this group, mostly resembling low-density polymorphonucleocytes (LD-PMN) (Fig 3N) that have been described in sepsis and systemic inflammatory response syndrome, and that might play multiple immunomodulatory activity, including the suppression of T cell responses^18^.

Collectively, these set of data suggest that in patients over 60 years old there is a specific immune signature characterized by suppression of T cell responses and deregulated innate immunity. In this regard, the increased level of circulating IL-15 in severe patients, having a longer hospitalization, might nourish the expansion of NK cell subsets in aged people as compared to the younger; on the other side, the prolonged exposure of NK cells to the circulating IL-15 might be responsible for the reduction of their cytolytic activity, potentially triggering a deregulated exhausted phenotype of these cells in the >60 group^19^. We also confirmed an exhausted makeup of both CD4^+^ and CD8^+^ T lymphocytes in older patients; this could represent an additional suppressive mechanism, feed by the peculiar cytokine milieu that contributed to the inadequate immune response against the SARS-CoV-2 virus in aged patients. Among all cytokines, we identified IL-27 as the circulating factor that showed the best correlation coefficient with age (R Parson=0.6097, Fig 2D), being also associated with HT and DS (Fig 3A). In COVID-19 patients, IL-27 might be involved in the up-regulation of inhibitory receptors such as TIM3 and in skewing T cell responses. Moreover, IL-27 might modulate T cell activity by inducing IL-10 production^20^, that we identified to be also increased in patients in relation to age and DS. In turn, IL-10 might feed immunosuppressive circuits orchestrated by LD-PMN, that were expanded in older COVID-19 patients, suggesting the possible targeting of immunosuppressive checkpoints for novel therapeutic approaches. Two other cytokines of the shared signature, TNF-α and CXCL8, were also suggestive of bolstered innate immune responses in older SARS-CoV-2 infected individuals. In line with this, anti-TNF trials have been recommended for COVID-19 patients who developed *acute respiratory distress syndrome* (ARDS) in the 2 days following hospital admission^21^. Consistently with available studies^22^, increased CXCL8 level in our cohort delineated a severe illness in patients and it might also represent a predictive biomarker of acute lung injury and ARDS in these patients^23^.

In conclusion, this study identified distinctive immunological features of COVID-19 patients that associate with age and are predictors of disease severity. Being our analysis performed at hospital admission, this study suggests novel markers that can be explored to identify novel criteria for COVID-19 patient stratification at hospital entry. In addition, the analyses revealed specific age-dependent immune signatures that may shed light on COVID-19 pathogenesis.

## Ethical commitment

Local ethics committee was notified about the study protocol. The study was performed according to the ethical guidelines of the Declaration of Helsinki (7th revision). All the patients gave their written informed consent and all analyses were carried out on anonymised data as required by the Italian Data Protection Code (Legislative Decree 196/2003) and the general authorization issued by the Data Protection Authority.

## Data Availability

The data supporting the findings of this study are available from the corresponding authors upon written request.

## Acknowledgements

We thank all the patients who participated to the study, the entire clinical staff of the Infectious Disease Unit of the Padova University Hospital. We also thank Euroimmun for IgG test screening and Prof. E. Bucci for the critical reading of the manuscript.

The study was funded by Fondazione Città della Speranza, grant number 20/02CoV, to AV.

## Author contributions

RA, RSR, FM performed all the experiments and analysis; NB provided technical support for sample processing; DA performed Milliplex Assay; SC and DM enrolled patients, analyzed demographic and clinical data and coordinated blood sampling; AC supervised all the clinical aspects of the study and contributed to the interpretation of the results; RA RSR FM AV and BM conceived the study; AV and BM discussed the results and wrote the manuscript.

## Figure legends

**Figure 1S.**
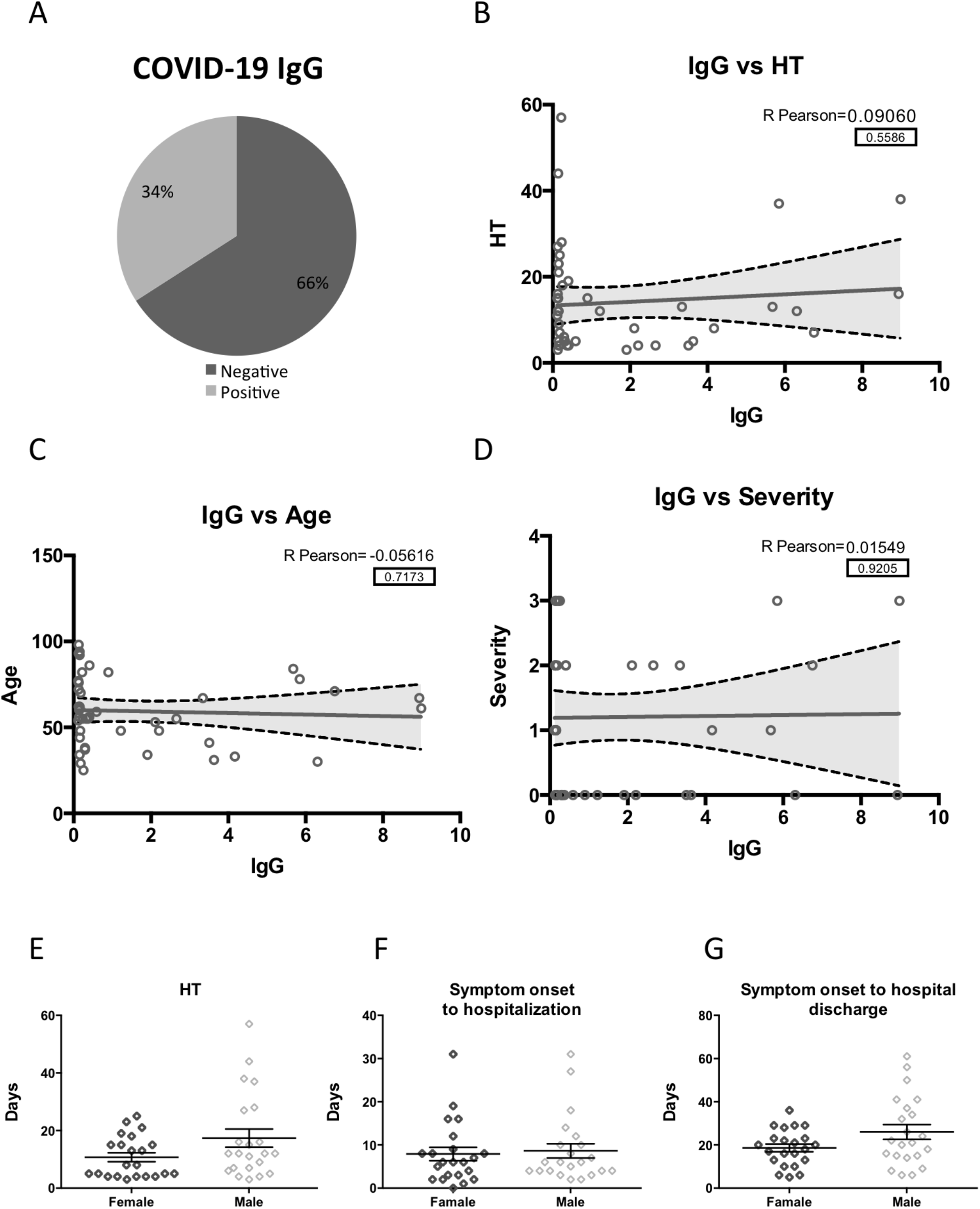
HT and DS are independent from IgG response at admission time and sex. Circle-graph showing the percentage of anti-SARS-CoV-2 (IgG) positive patients at HA **(A)**. Correlation analysis of SARS-CoV-2 -specific IgG with HT (**B**), age (**C**) and DS (**D**) measured by Person coefficient r (95% confidence interval) and two-tailed p-value analysis (indicated inside the square). Sex-matched analysis of HT (**E**), days from symptoms onset to HA (**F**) and days from symptom onset to hospital discharge) (**G**).

**Figure 2S.**
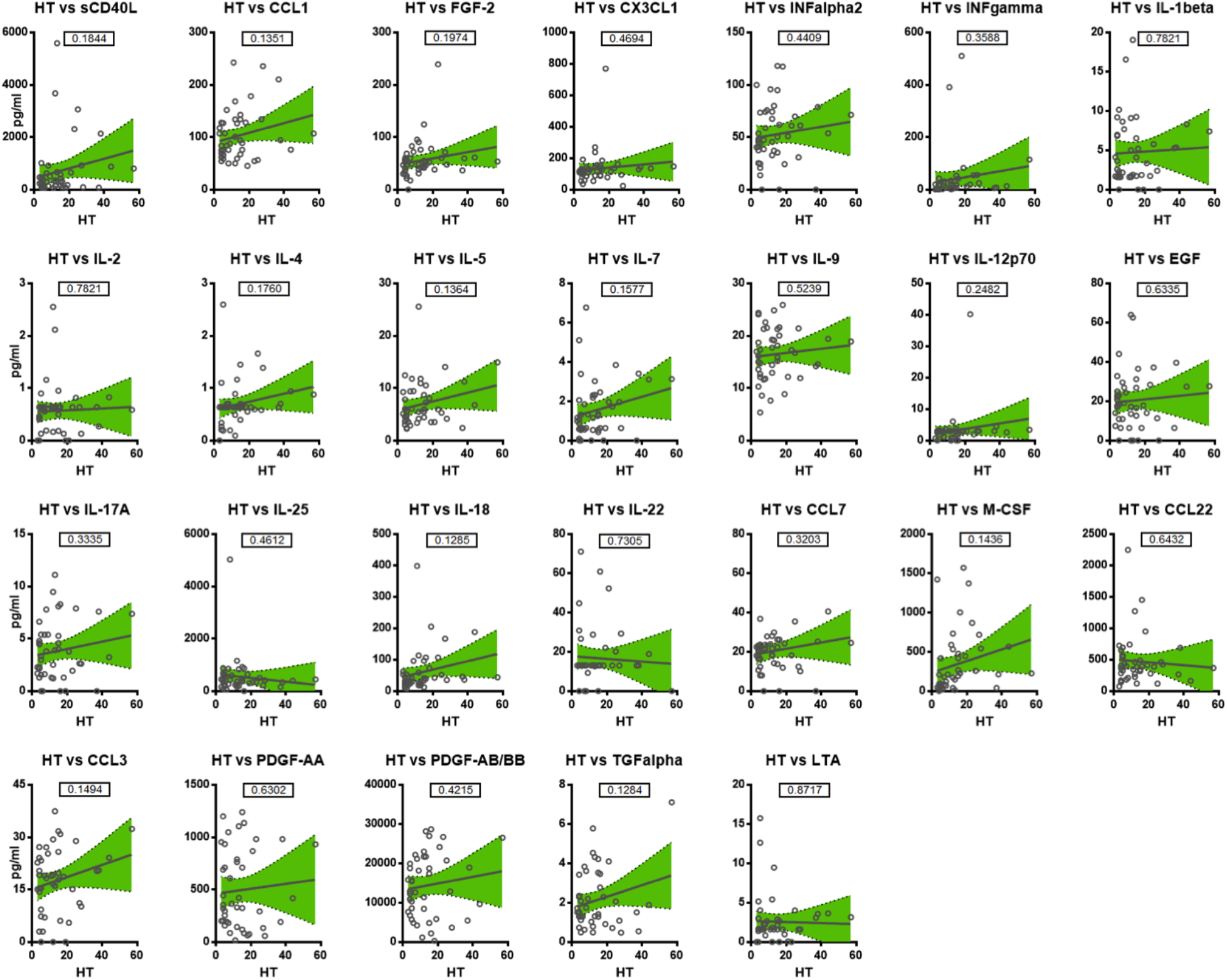
HT-cytokines. Correlation analysis of analytes and HT measured by Person coefficient r (95% confidence interval) and two-tailed p-value analysis (indicated inside the square).

**Figure 3S.**
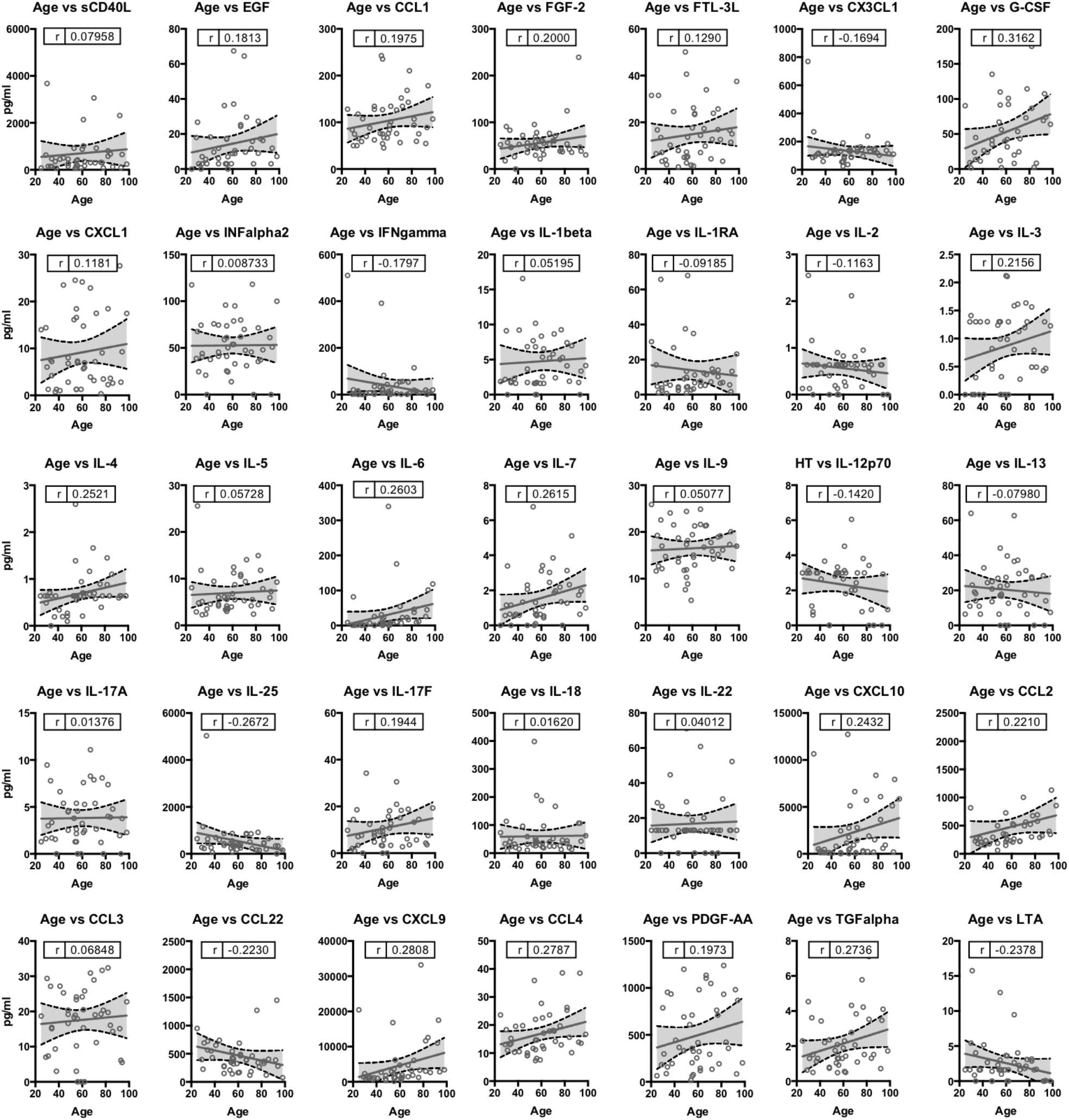
Age-cytokines. Correlation analysis of analytes and age measured by Person coefficient r (95% confidence interval) and two-tailed p-value analysis (indicated inside the square).

**Figure 4S.**
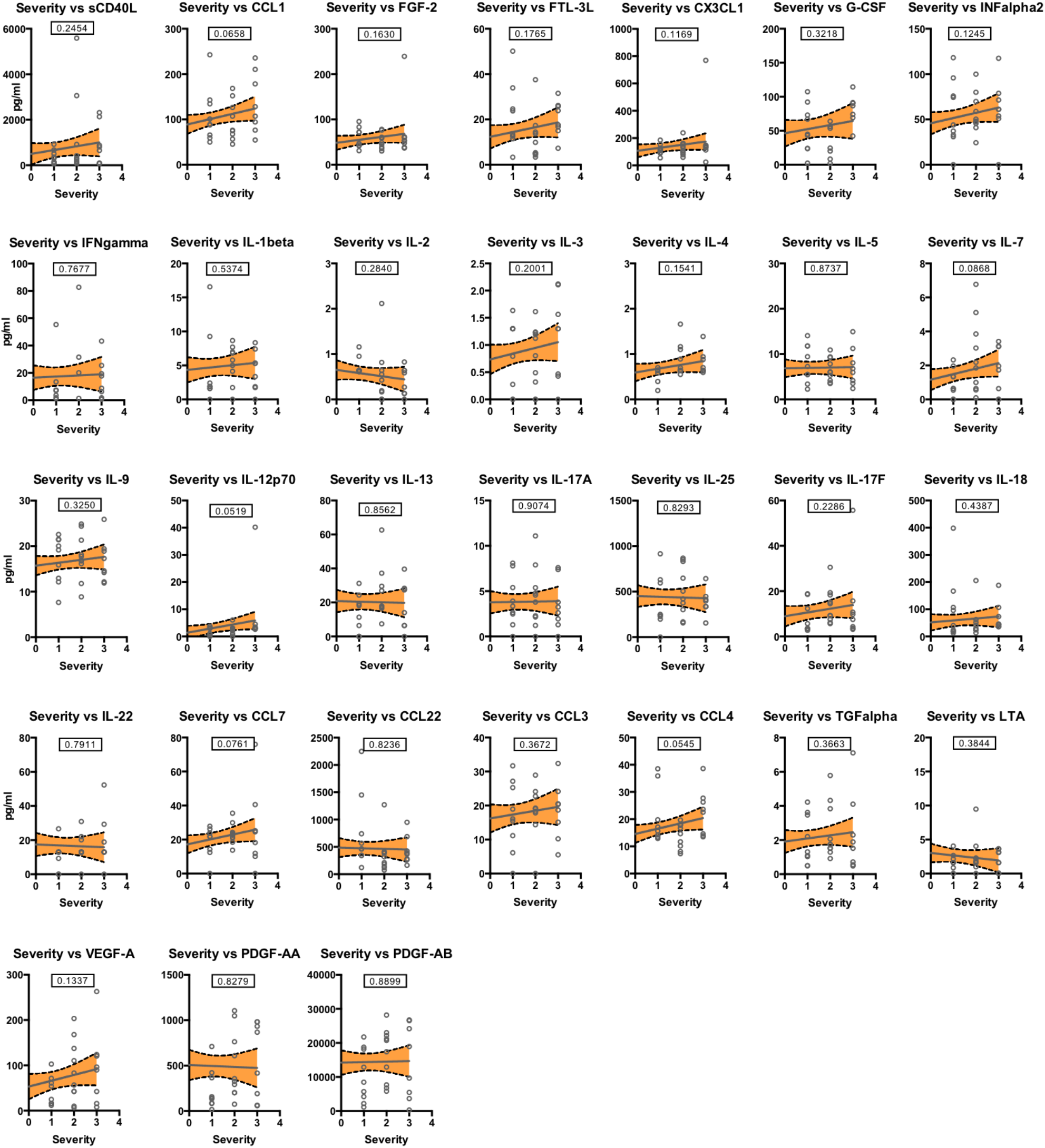
DS-cytokines. Correlation analysis of analytes and DS measured by Person coefficient r (95% confidence interval) and two-tailed p-value analysis (indicated inside the square).

**Figure 5S.**
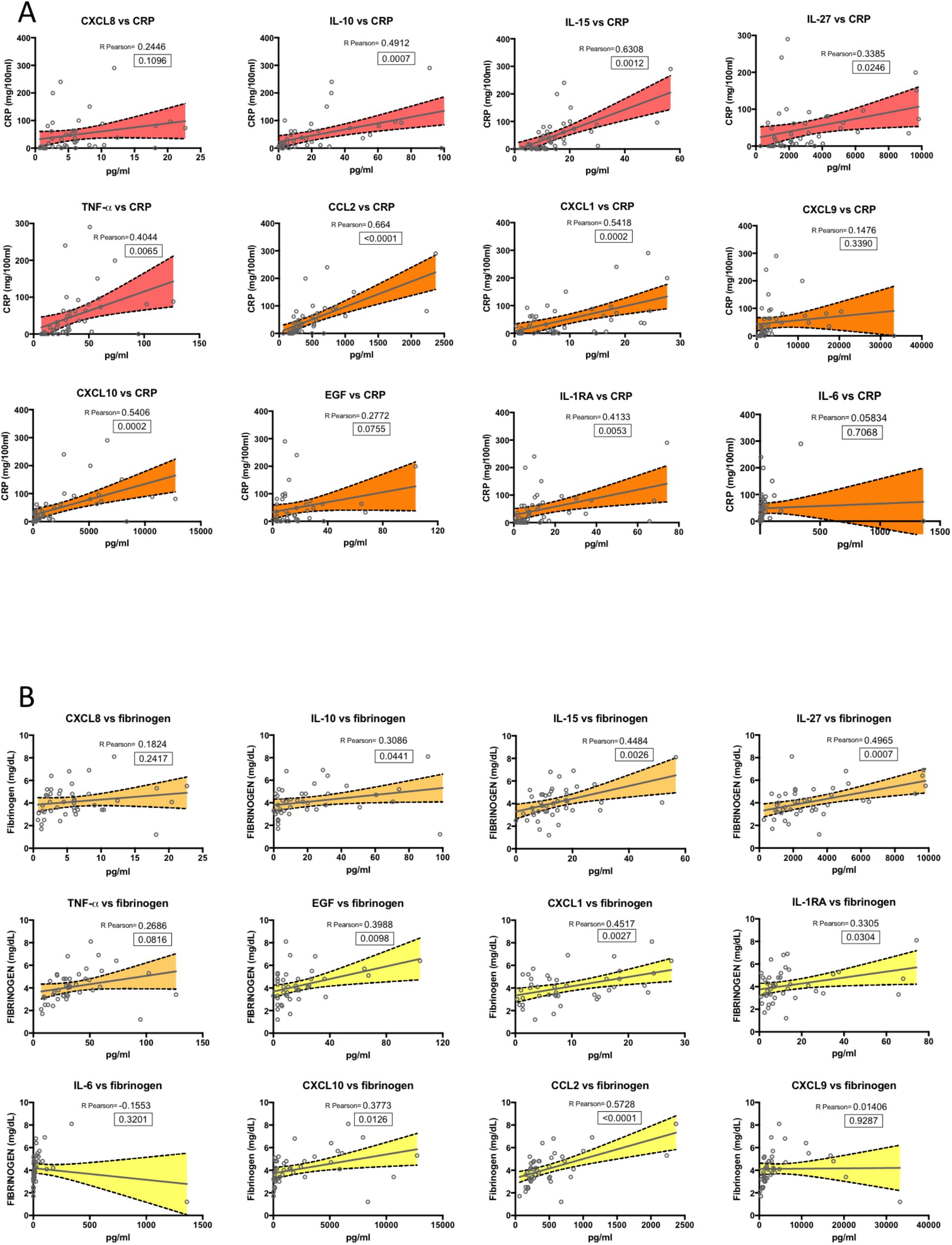
Clinical relevance of age-dependent and age-independent cytokines. Correlation analysis of age-dependent cytokines (*red*) and age-independent (*orange*) with CRP **(A**) and age-dependent cytokines (*light orange*) and age-independent (*yellow*) with fibrinogen **(B**) value in COVID-19 patients, measured by Person coefficient r (95% confidence interval) and two-tailed p-value analysis (indicated inside the square).

**Figure 6S.**
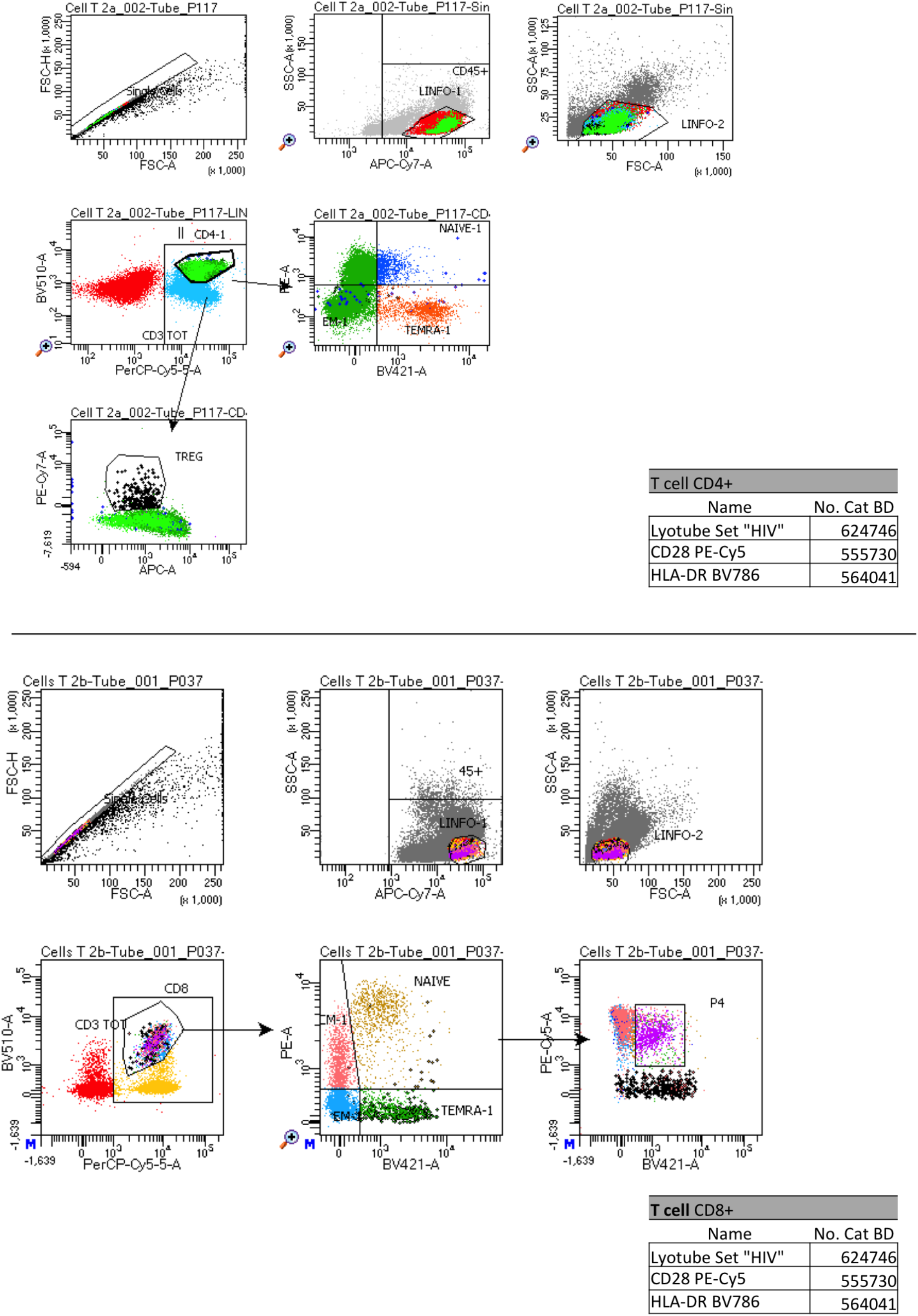
Gating strategy for the identification of T cell subsets.

**Figure 7S.**
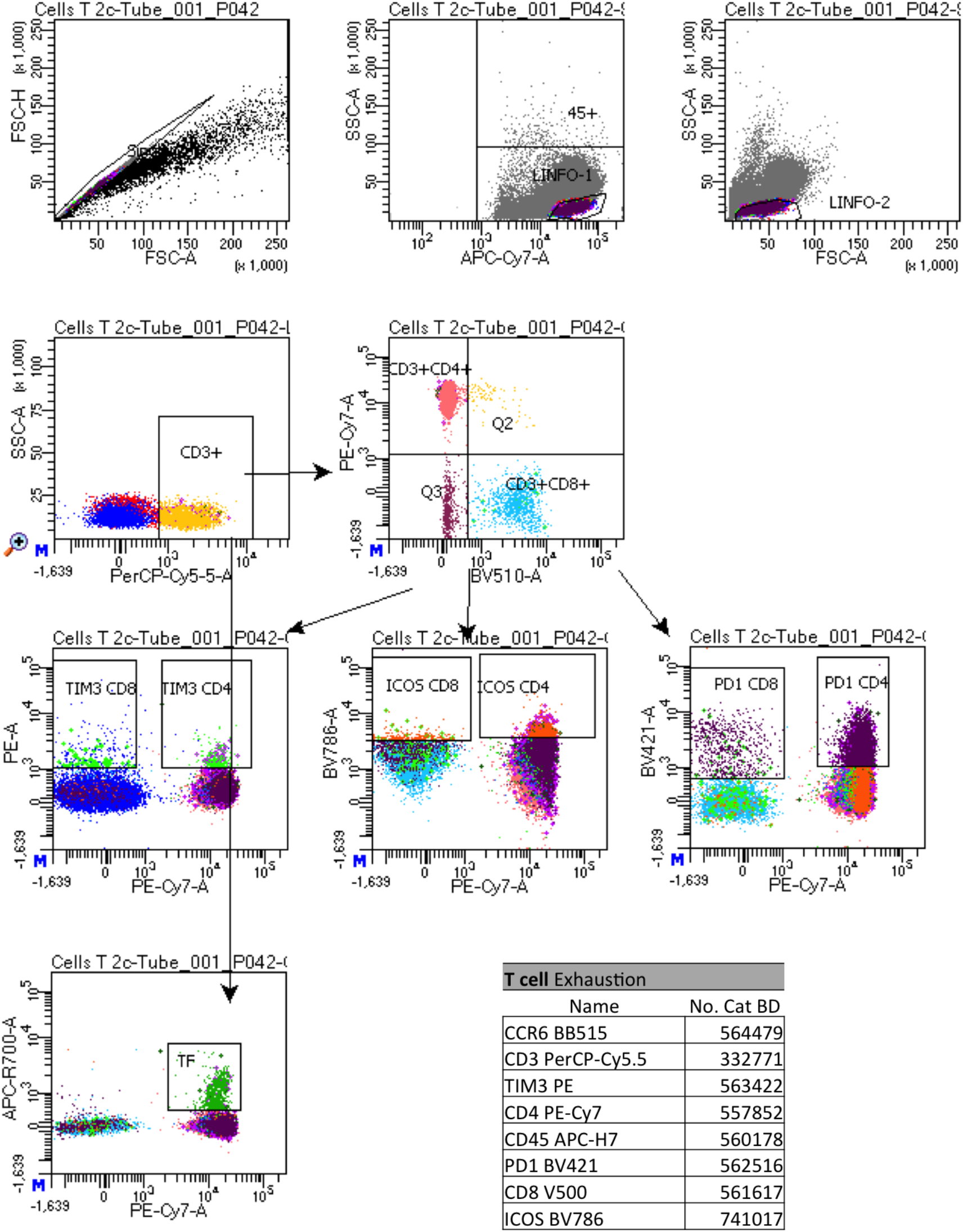
Gating strategy for the identification of T cell exhaustion.

**Figure 8S.**
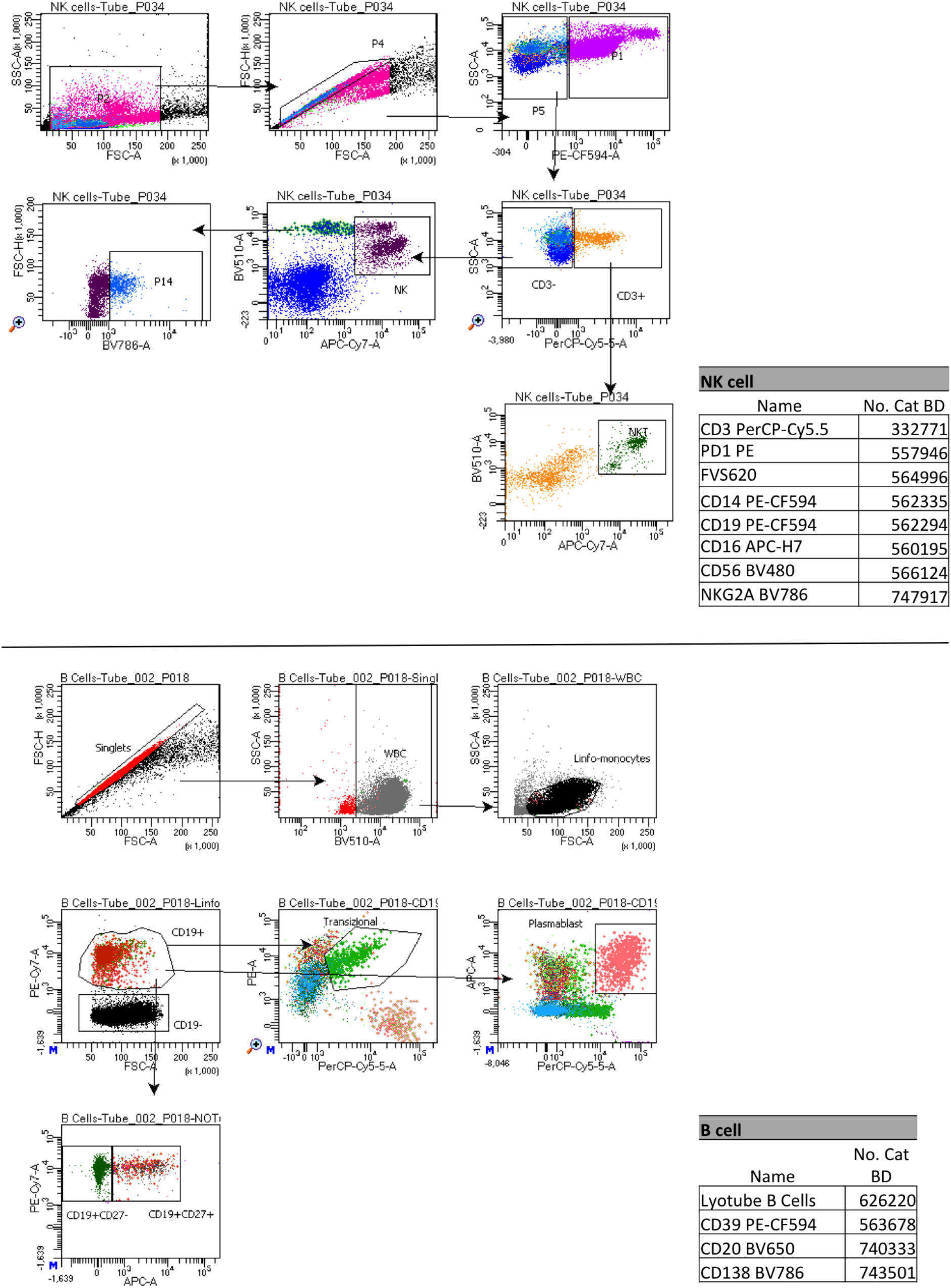
Gating strategy for the identification of NK and B cells.

**Figure 9S.**
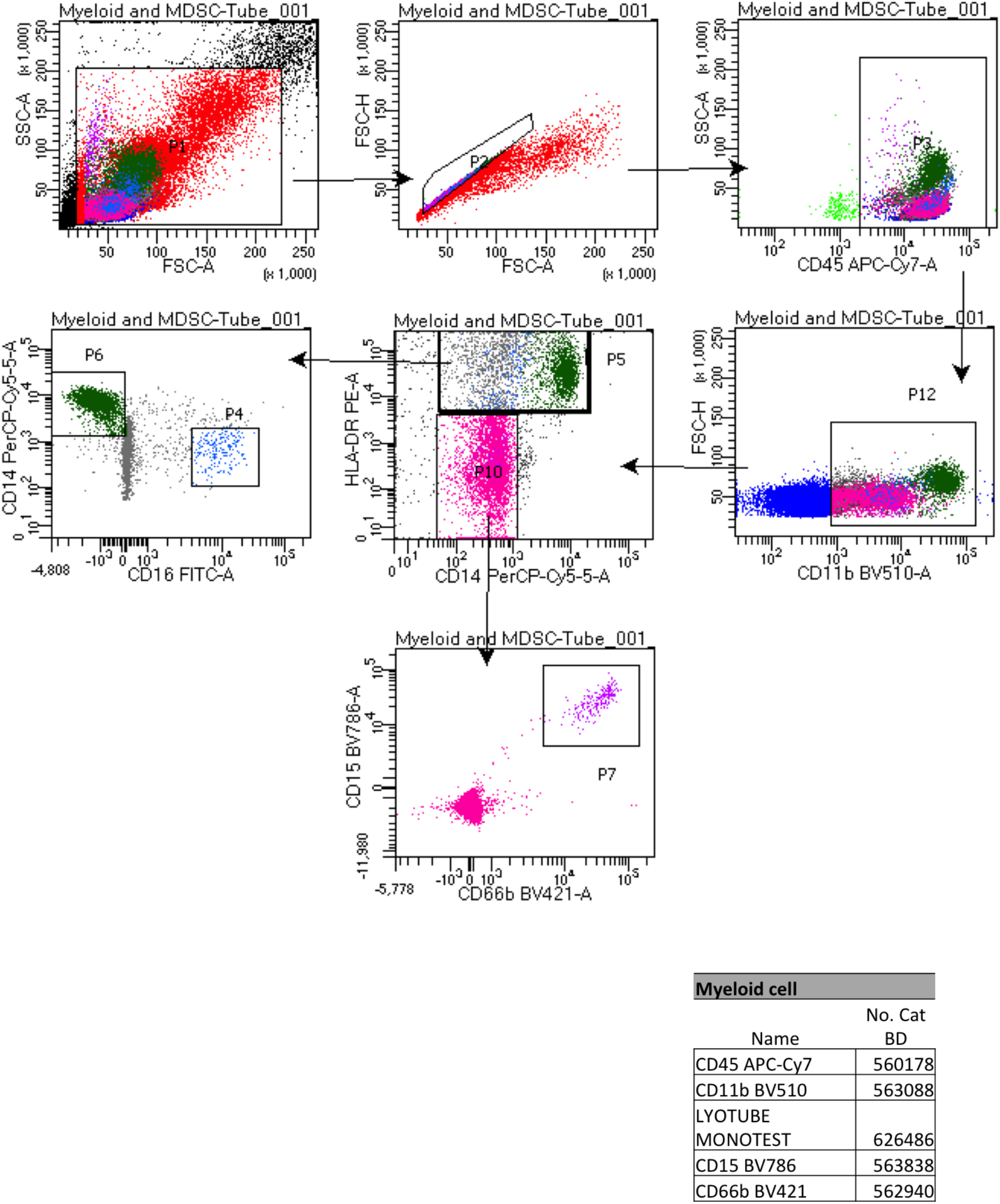
Gating strategy for the identification of myeloid cell subsets.

**Figure 10S.**
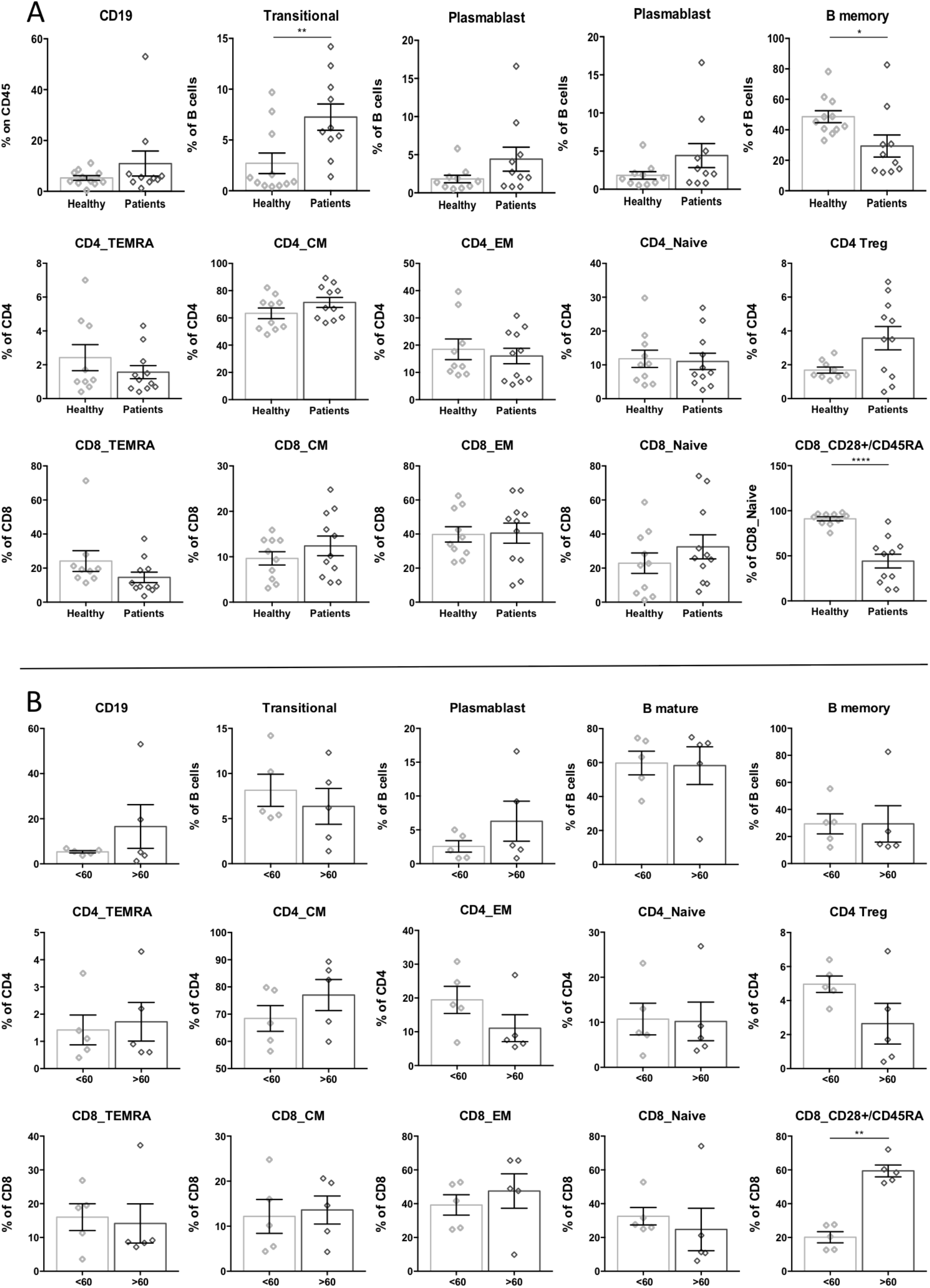
Immune cell population in age-matched Covid-19 patients. Percentage of immune cell populations assessed by FACS analysis in healthy age-matched controls and COVID-19 patients (**A**), and in younger (<60) or older (>60) Covid-19 patients (**B**).

## Methods

### Participants, study design and data collection

44 adult patients hospitalized at the COVID-19 center of the Infectious Diseases Division (IDD) of the University Hospital of Padua, Italy, between 9.04.2020 and 5.05.2020 were enrolled in the study. All patients were diagnosed with COVID-19 with SARS-CoV-2 infection confirmed by real-time reverse transcription polymerase chain reaction method (WHO guidelines). 12 SARS-CoV-2-negative age-matched participants were considered as a control group. All patients were classified into mild, moderate, severe and critical cases based on results from chest imaging, clinical examination, and symptoms (WHO guidelines). In addition, patients were stratified in two groups by fixing the age of 60 as the cut-off for patient grouping.

Demographic, clinical, laboratory data were extracted from paper and electronic medical records using a standardized data collection form. Laboratory data included: complete blood count, ESR, CRP, coagulation profile, serum biochemical tests. Both Chest X rays and CT scans were performed for all COVID19 patients.

### PBMC isolation

Peripheral Blood (PB) from enrolled controls and COVID-19 inpatients. PB was collected in EDTA tubes and stored at 4°C prior to processing for PBMC isolation and plasma collection. Peripheral blood mononuclear cells (PBMC) were isolated by density-gradient sedimentation using Ficoll-Paque PLUS (GE Healthcare, Germany) according to manufacturer’s protocol. Post-purification the isolated PBMC were cryopreserved in cell recovery media containing 10% DMSO (Gibco), supplemented with 10% heat inactivated HyClone™ Fetal Bovine Serum (FBS; GE Healthcare, Germany) and stored in liquid nitrogen. Plasma was then carefully removed from the 2/3 of the top layer using a sterile serological pipette until the mononuclear cell interphase, portioned and aliquots were stored at −80C until the analysis.

### Flow Cytometry

PBMC immune cell phenotyping was performed by custom set-up panels and BD Lyotubes by multiparametric FACS analysis. PBMC were thawed in RPMI 1640 with L-Glutamine (LONZA) medium with 2% FBS (GE Healthcare, Germany). 1×10^6^ PBMC were resuspended in 100 µl PBS with 2% FBS (FACS buffer) and stained with antibody cocktails at 4°C in the dark. Following surface staining, cells were washed with FACS buffer and fixed in FACS buffer/1%PFA. After two final washes, cells were resuspended in 200µl FACS buffer and acquired on a BD FACSCelesta™ Cell Analyzer (BD Biosciences, San Diego, CA). A list of antibodies used in the multicolor panels can be found in **Fig S6-S7-S8-S9**.

### Luminex assay

48 analytes (sCD40L, EGF, Eotaxin, FGF-2, Flt-3 ligand, CX3CL1, G-CSF, GM-CSF, CXCL1, IFNα2, IFNγ, IL-1α, IL-1β, IL-1ra, IL-2, IL-3, IL-4, IL-5, IL-6, IL-7, CXCL8, IL-9, IL-10, IL-12 (p40), IL-12 (p70), IL-13, IL-15, IL-17A, IL-17E/IL-25, IL-17F, IL-18, IL-22, IL-27, CXCL10, CCL2, CCL7, M-CSF, CCL22, CXCL9, CCL3, CCL4, PDGF-AA, PDGF-AB/BB, CCL5, TGF-α, TNF-α, TNF-β, VEGF-A) were analyzed by Luminex assay (Millipore, Billerica, USA) in the plasma from controls and 44 patients. The diluted standard and quality control were used according to the manufacture’s instruction. The plate was read on Luminex 200™. Analysis was performed using xPONENT 3.1 software.

### Immunoglobulin testing

SARS-CoV-2 (Spike S1domain epitope) IgG and IgA detection were performed by ELISA Assay Kit from Euroimmun according to the manufacture’s instruction. (https://www.euroimmun.it/coronavirus-2019-ncov/)

### Data Quantification and Statistical Analysis

Flow cytometry data were analyzed with BD FACSDiva™ (BD, Italy) and statistical analyses were done in Prism 8.4 (GraphPad, USA). Pearson correlation test was used to estimate the association between two quantitative variables *p*-value < 0.05 were considered statistically significant. Data plotted were expressed as Mean with Standard Error of Mean (SEM). Data distribution was assessment by D’agostino-Pearson and Shapiro-Wilk normality test. Unpaired Nonparametric *t*-test or Krustal-Wallis test followed by *post hoc* Dunn’s multiple comparations was used to compare differences between groups. Differences were considered statistically significant at confidence levels *P< 0.05 or **P< 0.01.

## References

1. Wilk, A. J. et al. A single-cell atlas of the peripheral immune response in patients with severe COVID-19. Nat. Med. (2020). doi:10.1038/s41591-020-0944-y

2. Banerjee, A. et al. Estimating excess 1-year mortality associated with the COVID-19 pandemic according to underlying conditions and age: a population-based cohort study. Lancet (2020). doi:10.1016/S0140-6736(20)30854-0

3. WHO. Clinical Management of COVID-19 - Interim Guidance. WHO (2020).

4. Bi, Q. et al. Epidemiology and transmission of COVID-19 in 391 cases and 1286 of their close contacts in Shenzhen, China: a retrospective cohort study. Lancet Infect. Dis. (2020). doi:10.1016/S1473-3099(20)30287-5

5. Qi, Z. & Yu, Y. Epidemiological Features of the 2019 Novel Coronavirus Outbreak in China. Curr. Top. Med. Chem. (2020). doi:10.2174/1568026620999200511094117

6. Sun, B. et al. Kinetics of SARS-CoV-2 specific IgM and IgG responses in COVID-19 patients. Emerg. Microbes Infect. (2020). doi:10.1080/22221751.2020.1762515

7. Phipps, W. S. et al. SARS-CoV-2 Antibody responses do not predict COVID-19 disease severity. medrxiv (2020). doi:10.1101/2020.05.15.20103580

8. Maleki Dana, P. et al. An Insight into the Sex Differences in COVID-19 Patients: What are the Possible Causes? Prehosp. Disaster Med. (2020). doi:10.1017/s1049023x20000837

9. Qin, C. et al. Dysregulation of immune response in patients with COVID-19 in Wuhan, China. Clin. Infect. Dis. (2020). doi:10.1093/cid/ciaa248

10. Schett, G., Sticherling, M. & Neurath, M. F. COVID-19: risk for cytokine targeting in chronic inflammatory diseases? Nature Reviews Immunology (2020). doi:10.1038/s41577-020-0312-7

11. Wan, S. et al. Characteristics of lymphocyte subsets and cytokines in peripheral blood of 123 hospitalized patients with 2019 novel coronavirus pneumonia (NCP). medRxiv (2020). doi:10.1101/2020.02.10.20021832

12. Chi, Y. et al. Serum Cytokine and Chemokine profile in Relation to the Severity of Coronavirus disease 2019 (COVID-19) in China. J. Infect. Dis. (2020). doi:10.1093/infdis/jiaa363

13. Huang, C. et al. Clinical features of patients infected with 2019 novel coronavirus in Wuhan, China. Lancet (2020). doi:10.1016/S0140-6736(20)30183-5

14. Dudley, J. P. & Lee, N. T. Disparities in Age-Specific Morbidity and Mortality from SARS-CoV-2 in China and the Republic of Korea. Clin. Infect. Dis. (2020). doi:10.1093/cid/ciaa354

15. et al. Age-dependent effects in the transmission and control of COVID-19 epidemics. Nat. Med. (2020). doi:10.1038/s41591-020-0962-9

16. Kang, S. J. & Jung, S. I. Age Related Morbidity and Mortality among Patients with COVID-19. Infect. Chemother. (2020).

17. Zheng, M. et al. Functional exhaustion of antiviral lymphocytes in COVID-19 patients. Cellular and Molecular Immunology (2020). doi:10.1038/s41423-020-0402-2

18. Silvestre-Roig, C., Fridlender, Z. G., Glogauer, M. & Scapini, P. Neutrophil Diversity in Health and Disease. Trends in Immunology (2019). doi:10.1016/j.it.2019.04.012

19. Felices, M. et al. Continuous treatment with IL-15 exhausts human NK cells via a metabolic defect. JCI insight (2018). doi:10.1172/jci.insight.96219

20. Murugaiyan, G. et al. IL-27 Is a Key Regulator of IL-10 and IL-17 Production by Human CD4 + T Cells. J. Immunol. (2009). doi:10.4049/jimmunol.0900568

21. Remuzzi, A. & Remuzzi, G. COVID-19 and Italy: what next? The Lancet (2020). doi:10.1016/S0140-6736(20)30627-9

22. Gong, J. et al. Correlation Analysis Between Disease Severity and Inflammation-related Parameters in Patients with COVID-19 Pneumonia. medRxiv (2020). doi:10.1101/2020.02.25.20025643

23. Isabel García-Laorden, M., Lorente, J. A., Flores, C., Slutsky, A. S. & Villar, J. Biomarkers for the acute respiratory distress syndrome: How to make the diagnosis more precise. Annals of Translational Medicine (2017). doi:10.21037/atm.2017.06.49

